# The Colorado Heart Failure Acuity Risk Model (CHARM) Score: A Mortality Risk Model for Waitlisted Cardiac Transplant Patients

**DOI:** 10.1101/2023.08.30.23294870

**Authors:** Rachel D. Murphy, Sarah Y. Park, Larry A. Allen, Amrut V. Ambardekar, Joseph C. Cleveland, Michael T. Cain, Bruce Kaplan, Jordan R.H. Hoffman, John S. Malamon

## Abstract

**Importance:** Although the Organ Procurement and Transplantation network provides structured policies and guidance for waitlisted cardiac transplant patients, the heart transplantation community lacks a mathematical model that can accurately estimate the short-term risk of death associated with being waitlisted. Importantly, the CHARM score provides a risk management and ranking system for patients based on a well-defined and sensitive medical urgency metric.

**Objective:** We had three primary objectives in completing this study. First, to increase relevance and applicability, we selected patient attributes that were clinically justified and readily available. Second, we designed and implemented an intuitive, formal system that accurately defined the relative risk of death while being waitlisted at 30-day, 90-day, and 1-year censoring periods. Third, we developed and validated a medical urgency metric that is intuitive, practical, and can be implemented nationally.

**Design:** We present a multivariable, prognostic model and risk management strategy for adult waitlisted heart transplant patients (N=1,965) from the Scientific Registry of Transplant Recipients (SRTR) database that were waitlisted from January 1, 2008, to September 2, 2022. To independently validate each model, we randomly split this cohort into a discovery set (N=1,174) and validation set (N=784). Twelve independent patient attributes were selected, and three linear regression formulas were derived to estimate and rank the relative risk of dying while waitlisted. Four independent validation methods were used to measure each model’s performance as a classifier and ranking system.

**Setting:** The United States

**Participants:** This cohort (N=1,965) consisted of adult heart transplant candidates without missing laboratory data who were placed on the waitlist from January 1, 2008, to September 2, 2022. Patients listed for multi-organ transplantation were excluded.

Patients with missing laboratory data were analyzed independently.

**Exposures:** The short-term risk of death remaining on the heart transplant waitlist.

**Main Outcomes and Measures:** The primary outcome of this study was the design, development, and validation of a formal risk management system for waitlisted heart transplant candidates experiencing end-stage heart failure. We derived three linear regression formulas and calibrated a seven-tiered risk index to accurately rank patients who were more likely to die on the waitlist at 30-day (30D), 90-day (90D), and 1-year (1Y) censoring periods. Four independent validation methods were used to measure each model’s classification and ranking performance.

**Results:** Using six interaction terms, we applied the 5-fold cross-validation procedure to the CHARM to discover an area under the ROC curve of 96.4%, 90.4.%, and 78% for the 30D, 90D, and 1Y models, respectively. The mean positive predictive values of the tiered risk system were 99.2% (30D), 94.1% (90D), and 88% (1Y). Risk indices for all three models were >99% correlated to the observed mortality rate across the seven tiers for the 30D, 90D, and 1Y models.

**Conclusions and Relevance:** We designed, implemented, and validated an intuitive and formal risk scoring and ranking system which is ideal for prioritizing waitlisted heart failure patients based on a well-defined medical urgency metric. The CHARM score provides extreme sensitivity in predicting short-term mortality outcomes. The CHARM score is extensible to larger patient populations experiencing end-stage heart failure.

**KEY POINTS:** *Question:* Can pre-operative patient characteristics be used to develop a formal system to accurately estimate, rank, and predict the relative short-term mortality of waitlisted heart transplant patients?

*Findings:* Using twelve patient attributes, we derived three linear regression equations to accurately predict the 30-day, 90-day, and 1-year mortality of waitlisted heart transplant patients. We developed and calibrated a seven-tiered risk index for each model that was 99% correlated to the observed mortality rate. Using several independent validation methods, we achieved extreme sensitivity (>98%) in ordinally ranking patient groups who were more likely to survive 30 days on the waitlist. Model performance was measured using the area under the receiver operating characteristic (ROC) curve. Using six interaction terms, the area under the ROC curve was 96.4% (30-day), 90.4% (90-day), and 78% (1-year).

*Meaning:* Our models accurately discriminate among patient subgroups who are more likely to die while waitlisted. Because our tiered ranking system is simple, extremely sensitive, and well calibrated, it is ideal for prioritizing waitlisted heart transplant patients based on a well-defined medical urgency score. These models are generalized and therefore extensible to defining medical urgency in larger patient populations experiencing end-stage heart failure.

## 1. INTRODUCTION

The Organ Procurement and Transplantation Network (OPTN) has worked to develop a cardiac transplant allocation system that distributes donor hearts to the most critically ill patients^1^. The 2018 policy revision created a 6-tiered “status”-based stratification system that attempted to encompass the increasing complexity of managing critical cardiac illness. However, this policy has been critiqued for its subjectivity and heterogeneity in accurately discriminating patient risk while on the waitlist. For example, a large percentage of transplant candidates are now stratified to Status 1 or Status 2 by exception rather than by the standard criteria, which can be modified by a physician’s practice^2^. Importantly, studies have demonstrated that the 2018 policy revision is associated with a significant reduction in post-transplant survival^3,4^. A formal prognostic model that accurately stratifies waitlisted end-stage heart failure (HF) patients based on medical urgency is nonexistent. The need for such a model was recently highlighted by Pelzer *et al.*, who determined the current allocation system had only a moderate ability to successfully rank transplant candidates according to medical urgency^5^.

While useful prognostic tools for patients with HF have been introduced, such as the Heart Failure Survival Score^6^ and the Seattle Heart Failure Model^7^, they have not accurately predicted waitlist mortality^8^. The Ottawa Heart Failure Risk Scale provides a risk stratification tool designed for acute heart failure patients in emergency departments^9^. However, this score has not been applied to or validated against a waitlisted heart transplant patient population. Similarly, the Meta-Analysis Global Group in Chronic Heart Failure (MAGGIC) risk score performs well for HF patients with preserved ejection fraction, but it has not been externally validated for reduced ejection fraction, which would exclude most of the population on a transplant list^10^.

Predictive models have been successfully implemented in other organ transplant systems. The Model for End-stage Liver Disease including Sodium or MELD-Na score for liver transplantation accurately predicts 90-day patient waitlist mortality and is the most significant metric in liver allocation^11^. A similar model is desperately needed in cardiac transplantation to accurately prioritize the most critically ill patients. We used the Scientific Registry of Transplant Recipients (SRTR) database to develop and validate three predictive mortality models for waitlisted patients with end-stage HF. Our models utilize objective physiological data to determine the most urgent heart transplant candidates and stratify their relative risk of waitlist mortality at 30 days, 90 days, and 1 year.

## 2. METHODS

### 2.1 Data sources

This study utilized retrospective data from the SRTR. The SRTR system includes data on all donors, waitlisted candidates, and transplant recipients in the US, submitted by the members of OPTN^12^. The Health Resources and Services Administration, U.S. Department of Health and Human Services provides oversight to the activities of the OPTN and SRTR contractors. The data reported here have been supplied by the Hennepin Healthcare Research Institute as the contractor for SRTR. The interpretation and reporting of these data are the responsibility of the author(s) and in no way should be seen as an official policy of or interpretation by the SRTR or the U.S. Government.

### 2.2 Study population

The study population included adult waitlisted cardiac transplant patients with complete laboratory results who registered for a single-organ heart transplant (N=1,965) between January 1, 2008, and September 2, 2022. **Supplemental Figure 1** provides a participant workflow diagram with the number of participants retained for each exclusion criterion. All steps in this analysis were conducted for 30-day (30D), 90-day (90D), and 1-year (1Y) all-cause mortality. Survival times for waitlisted candidates started at the date of listing and were censored at the date of death and upon removal from the waitlist. The population was randomly split (60%/40%) into two cohorts, a discovery set (N=1,179) and a validation set (N=786). The discovery set was used for variable selection, the generation of three linear regression equations, and the creation of a tiered risk index. The validation set was used to independently evaluate each model’s performance in predicting mortality outcomes and ranking patients’ relative risk of death.

### 2.3 Study Ethics

Informed consent was obtained for all study participants. This study was reviewed by an ethical committee (Colorado Multiple Institutions Review Board) and was determined to be non-human subjects research. This study was not grant funded. The authors do not have any conflicts of interest to disclose. All authors reviewed the results and approved the final manuscript.

### 2.4 Statistical approach

#### 2.4.1 Missingness and Sensitivity Analysis

We compared the patient populations with some missing laboratory data (N=20,991) to those without missing data (N=1,965). For continuous variables, two-way ANOVA was used to test the observed differences in patient characteristics and the twelve independent variables used in this study. The chi-squared test was used to measure the significance of categorical and indicator (binary) variables. Missing data were significant in the laboratory variables; therefore, we analyzed missingness patterns and performed three related sensitivity analyses using: 1) participants without missing data (N=1,965); 2) all participants with missing laboratory data (N=22,949); and 3) all participants with missing laboratory values imputed (N=22,949). The Multivariate Imputation by Chain Variables or MICE algorithm was used to impute missing values.

#### 2.4.2 Clinical Variable Selection and Importance

For clinical applicability, we selected twelve patient variables that were readily available and clinically justified. These twelve variables should also exhibit discriminating power and low collinearity. To measure collinearity, we calculated the Pearson’s cross-correlation coefficient (R) and variance inflation factor (VIF) for all twelve independent variables. Low collinearity was defined as two independent variables with an R value less than 0.7. The VIF is used to determine the correlation between independent variables in a logistic regression model. A VIF of 1 provides no correlation, whereas values above 2.5 indicate considerable multicollinearity^13^. Using the discovery set, we measured and ranked the relative importance of each patient variable via the unsupervised Random Forest (RF) algorithm^14^ with the number of days survived as the outcome. The RF algorithm provided an orthogonal method for comparing the relative effect of each independent variable used in these models.

#### 2.4.3. Independent Variable Definitions

Life Support refers to any waitlisted heart transplant recipient who received any cardiac support prior to transplant including intravenous (IV) inotropic infusion, or any circulatory support device such as right ventricular assist devices (RVAD), left ventricular assist devices (LVAD), total artificial hearts (TAH), extracorporeal membrane oxygenation (ECMO), or intra-aortic balloon pumps (IABP)^15^. The Ventilator variable refers to the need for any mechanical ventilation prior to transplant. The use of mechanical ventilators has been shown to significantly increase patients’ risk of death post-transplant^16^. The Prior Cardiac Surgery indicator variable includes patients with any history of cardiac surgery.

#### 2.4.4 Regression Formulas for Calculating Patient Risk Scores

We set out to develop a short-term prognostic model to formally define a patient risk score also known as the CHARM score based on a patient’s estimated likelihood of death while waitlisted. Thus, we constructed three linear regression equations using the logistic link function for 30D, 90D, and 1Y mortality outcomes (**Equations 1-3**). All laboratory values were transformed to the logarithmic scale prior to calculation. We calculated CHARM scores for all patients in the discovery set to develop these equations. For consistency across the models, we used **Equation 4** to normalize CHARM scores to range from 0 to 1.

#### 2.4.5 Model Calibration and the Tiered Risk Index

A well calibrated ranking system is required to accurately estimate the relative medical urgency of waitlisted heart transplant patients. Thus, we developed a seven-tiered risk index system based on the CHARM score which ranges from one to seven. We then fit the risk tiers to the observed mortality rate to maximize the statistical discrimination in ranking the relative mortality likelihood observed in these patient subgroups or indices. We defined tiered risk thresholds and calibrated each regression model by maximizing the goodness-of-fit between the observed mortality and the risk indices, which are positively correlated. Here, we calculated the goodness-of-fit (R) using linear regression.

#### 2.4.6 Validating Dichotomous Outcomes using Logistic Regression

We measured each model’s performance in predicting mortality events at three censoring periods using 5-fold cross-validation (in-sample) and supervised sample hold-out validation (out-sample) using the validation set. We evaluated both additive models and those using interaction terms. Age, Albumin, Creatinine, Circulatory Support, Previous Transplant, and Prior Cardiac Surgery were the interaction terms. For sample hold-out validation, we used the supervised Random Forest classification method^14^ with 70 trees. Priors were calculated using the discovery set and predictions were independently validated using the validation set. Finally, we calculated the area under the ROC curve^17^ for all three models.

#### 2.4.7 Validating the Seven-Tiered Risk Index System

To validate our risk classification system using a univariate test, we leveraged the Cox proportional-hazard regression (CPHR) method^18^ to measure tier performance in terms of survival time. Here the unit of measurement was the concordance index^19^. To further evaluate the performance of the tiered risk system, we used **Equation 5** to calculate the rank precision or the positive predictive value (PPV) for each risk tier as compared to all others. A true positive (TP) occurred when a patient with a lower risk index outlived a patient in any higher tier. A false positive (FP) occurred when a patient with a higher risk index outlived a patient in any lower tier^20^. All analyses were performed using the R statistical language version 4.1.2^21^.

## 3. RESULTS

### 3.1 Population Characteristics

The discovery, validation, and total patient population characteristics are depicted in **Table 1**. The mean (SD) age of the 1,965 study participants with no missing laboratory data was 48.5 (14.5) years; 1,432 (73.1%) participants were male, 1,402 (71.6%) were White/Caucasian, 374 (19.1%) were Black or African American, 104 (5.3%) were Hispanic or Latino, 56 (2.9%) were Asian, 9 (0.5%) were Native Hawaiian or Other Pacific Islander, and 6 (0.3%) were American Indian or Alaska Native. In summary, there were no statistically significant differences in the patient characteristics or independent variables when comparing the discovery set to the validation set.

**Table 1.** Patient Characteristics and Laboratory Values for Waitlisted Heart Transplant Patients, 2008 to 2022.

|  |  |  |  |  |  |
| --- | --- | --- | --- | --- | --- |
|  |  | **Discovery**<br>**(N=1,179)** | **Validation**<br>**(N=786)** | **All (N=1,965)** | **p-value** |
| **Age** |  |  |  |  |  |
|  | Mean (SD) | 48.9 (14.6) | 47.8 (14.2) | 48.5 (14.5) | 0.243 |
|  | Median [Min, Max] | 53.0 [18.0, 74.0] | 50.0 [18.0, 73.0] | 52.0 [18.0, 74.0] |  |
| **Sex** |  |  |  |  |  |
|  | Female | 312 (26.6%) | 214 (27.3%) | 526 (26.9%) | 0.94 |
|  | Male | 862 (73.4%) | 570 (72.7%) | 1432 (73.1%) |  |
| **Race** |  |  |  |  |  |
|  | Caucasian | 851 (72.5%) | 551 (70.3%) | 1402 (71.6%) | 0.995 |
|  | Hispanic/Latino | 57 (4.9%) | 47 (6.0%) | 104 (5.3%) |  |
|  | Black or African<br>American | 222 (18.9%) | 152 (19.4%) | 374 (19.1%) |  |
|  | Asian | 31 (2.6%) | 25 (3.2%) | 56 (2.9%) |  |
|  | American Indian or<br>Alaska Native | 4 (0.3%) | 2 (0.3%) | 6 (0.3%) |  |
|  | Native Hawaiian or<br>Pacific Islander | 5 (0.4%) | 4 (0.5%) | 9 (0.5%) |  |
| **Ethnicity** |  |  |  |  |  |
|  | Latino | 59 (5.0%) | 47 (6.0%) | 106 (5.4%) | 0.65 |
|  | Non-Latino or<br>unknown | 1115 (95.0%) | 737 (94.0%) | 1852 (94.6%) |  |
| **Education** |  |  |  |  |  |
|  | High School (9-12) | 394 (33.6%) | 252 (32.1%) | 646 (33.0%) | 0.994 |
|  | Attended<br>College/Technical<br>School | 324 (27.6%) | 225 (28.7%) | 549 (28.0%) |  |
|  | Associate/Bachelor<br>Degree | 251 (21.4%) | 164 (20.9%) | 415 (21.2%) |  |
|  | Post-college<br>Graduate Degree | 114 (9.7%) | 88 (11.2%) | 202 (10.3%) |  |
|  | Grade School (0-8) | 21 (1.8%) | 15 (1.9%) | 36 (1.8%) |  |
|  | None | 70 (6.0%) | 40 (5.1%) | 110 (5.6%) |  |

| **Independent Variables** |  |  |  |  |  |
| --- | --- | --- | --- | --- | --- |
| **Albumin** |  |  |  |  |  |
|  | Mean (SD) | 3.59 (0.578) | 3.59 (0.611) | 3.59 (0.591) | 0.991 |
|  | Median [Min, Max] | 3.60 [1.70, 6.40] | 3.60 [0.7, 6.50] | 3.60 [0.7, 6.50] |  |
| **AST** |  |  |  |  |  |
|  | Mean (SD) | 39.4 (74.0) | 39.8 (83.1) | 39.6 (77.7) | 0.993 |
|  | Median [Min, Max] | 27.0 [0.1, 1640] | 26.0 [0.1, 1810] | 26.5 [0.1, 1810] |  |
| **Bilirubin** |  |  |  |  |  |
|  | Mean (SD) | 0.975 (1.83) | 1.09 (1.77) | 1.02 (1.80) | 0.377 |
|  | Median [Min, Max] | 0.7 [0.1, 40.6] | 0.7 [0.1, 34.7] | 0.7 [0.1, 40.6] |  |
| **BNP** |  |  |  |  |  |
|  | Mean (SD) | 1690 (2280) | 1560 (2180) | 1640 (2240) | 0.478 |
|  | Median [Min, Max] | 776 [0, 10000] | 696 [5, 10000] | 741 [0, 10000] |  |
| **Cardiac Surgery** |  |  |  |  |  |
|  | Yes | 506 (43.1%) | 332 (42.3%) | 838 (42.8%) | 0.947 |
|  | No | 668 (56.9%) | 452 (57.7%) | 1120 (57.2%) |  |
| **Creatinine** |  |  |  |  |  |
|  | Mean (SD) | 1.49 (1.16) | 1.53 (1.14) | 1.51 (1.15) | 0.725 |
|  | Median [Min, Max] | 1.24 [0.2, 16.1] | 1.25 [0.14, 10.8] | 1.25 [0.14, 16.1] |  |
| **Circulatory Support** |  |  |  |  |  |
|  | Yes | 436 (37.1%) | 296 (37.8%) | 732 (37.4%) | 0.962 |
|  | No | 738 (62.9%) | 488 (62.2%) | 1226 (62.6%) |  |
| **Previous Transplant** |  |  |  |  |  |
|  | Yes | 97 (8.3%) | 75 (9.6%) | 172 (8.8%) | 0.607 |
|  | No | 1077 (91.7%) | 709 (90.4%) | 1786 (91.2%) |  |
| **Sodium** |  |  |  |  |  |
|  | Mean (SD) | 135 (4.39) | 135 (4.63) | 135 (4.50) | 0.13 |
|  | Median [Min, Max] | 136 [117, 150] | 136 [109, 149] | 136 [109, 150] |  |
| **Ventilator** |  |  |  |  |  |
|  | Yes | 18 (1.5%) | 5 (0.6%) | 23 (1.2%) | 0.197 |
|  | No | 1156 (98.5%) | 779 (99.4%) | 1935 (98.8%) |  |
| **Waitlist Days** |  |  |  |  |  |
|  | Mean (SD) | 260 (388) | 268 (412) | 263 (398) | 0.899 |
|  | Median [Min, Max] | 110 [1.00, 4090] | 119 [1.00, 3370] | 113 [1.00, 4090] |  |
| *AST=aspartate aminotransferase; BNP=Brain natriuretic peptide* |  |  |  |  |  |

### 3.2. Missingness and Sensitivity Analysis

**Supplemental Table 1** summarizes the missing data for the twelve independent variables used in this model. More than 90% of total waitlisted patients were missing laboratory data. **Supplemental Table 2** provides the patient population characteristics for those with some missing data (N=20,991) and those without missing data (N=1,965). On average, patients without missing data were younger and more likely to be female or Caucasian. Statistically significant differences between the two cohorts were observed in five of the twelve independent variables. **Supplemental Figure 2** provides a missing value map for all independent variables. In summary, Age, Circulatory Support, Previous Transplant, Sodium, and Ventilator were missing at random. The other seven variables were missing completely at random^22^. **Supplemental Table 3** provides the AUC of the ROC for participants without missing data, with missing data, and with missing values imputed.

### 3.3 Variable Selection and Importance

**Supplemental Table 4** provides the relative variable importance in the RF model, measured by the Gini Index, a statistic dispersion value notated as the Increase in Node Purity (IncNodePurity). These data were normalized using **Equation 4** and are useful for ranking the relative importance of each independent variable. All logistic regression coefficients, p-values, and VIF values are provided in **Supplemental Table 5**. The correlation coefficients of the independent variables were below the absolute value of 0.21, and VIF values ranged from 1 to 1.532 (**Figure 1**). No significant co-linearity was observed in the variables or models.

**Figure 1.**
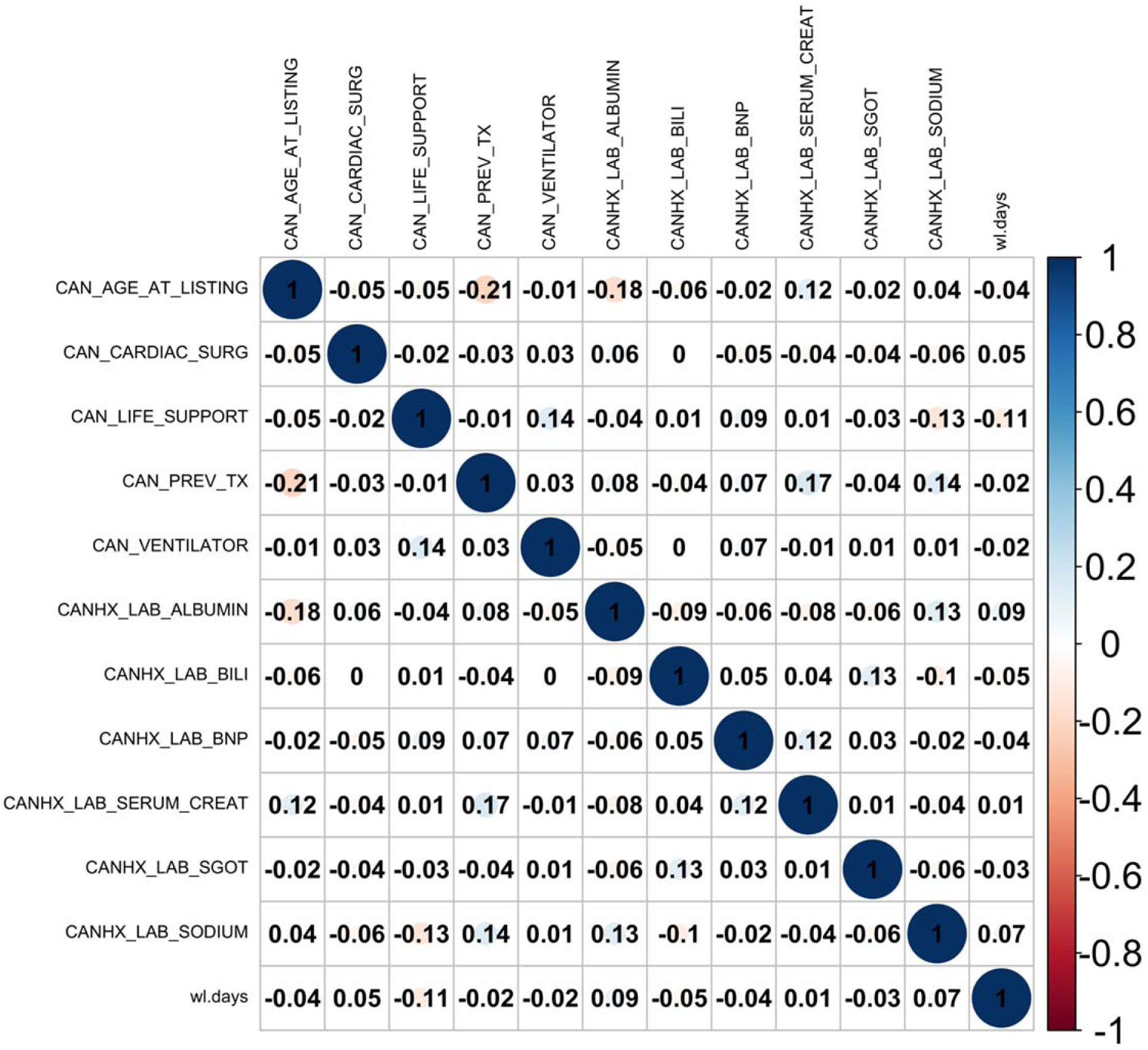
Correlation Heatmap of Independent Variables. Pearson’s correlation coefficients were calculated using all patients (N=1,965) and independent variables (N=12) used to construct the three models (30D, 90D, and 1Y). This is used to provide a measure of collinearity. Blue indicates a positive correlation, and red indicates a negative correlation. The color saturation and circle area increase as the correlation coefficients increase in magnitude.

### 3.4 Mathematical Formulas

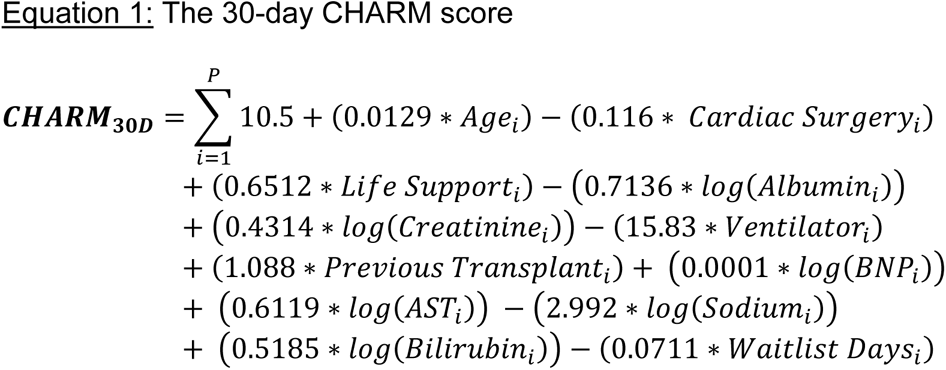

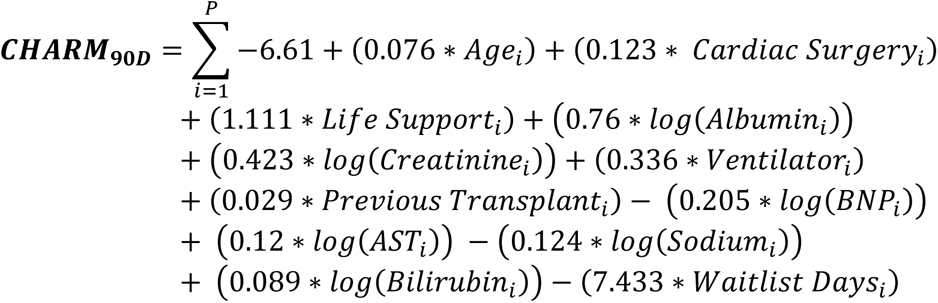

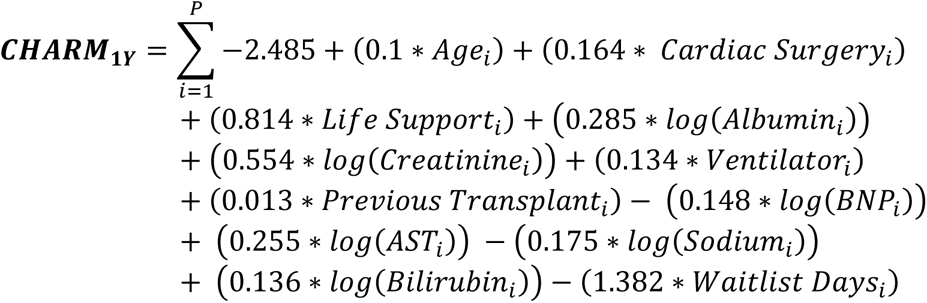

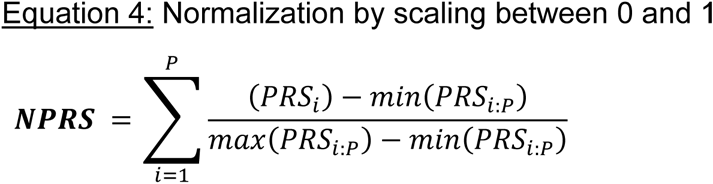

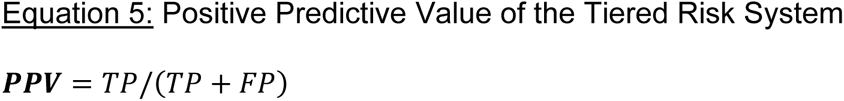

### 3.4 Model Calibration and the Tiered Risk Index

The observed mortality rate is presented in **Figure 2** per risk index as a function of the CHARM score**. Supplemental Figures 3-5** provide the CHARM score distributions for each model. For the 90-day model, Risk Index (RI) 1 had an observed mortality rate of 1.52%, RI 2 had a mortality rate of 3.54%, RI 3 had a mortality rate of 7%, RI 4 had a mortality rate of 11.25%, RI 5 had a mortality rate of 13.5%, RI 6 had a mortality rate of 20.8%, and RI 7 had a mortality rate of 26.5%. The risk indices were >99% correlated to the observed mortality rate across the seven tiers for all three models.

**Figure 2.**
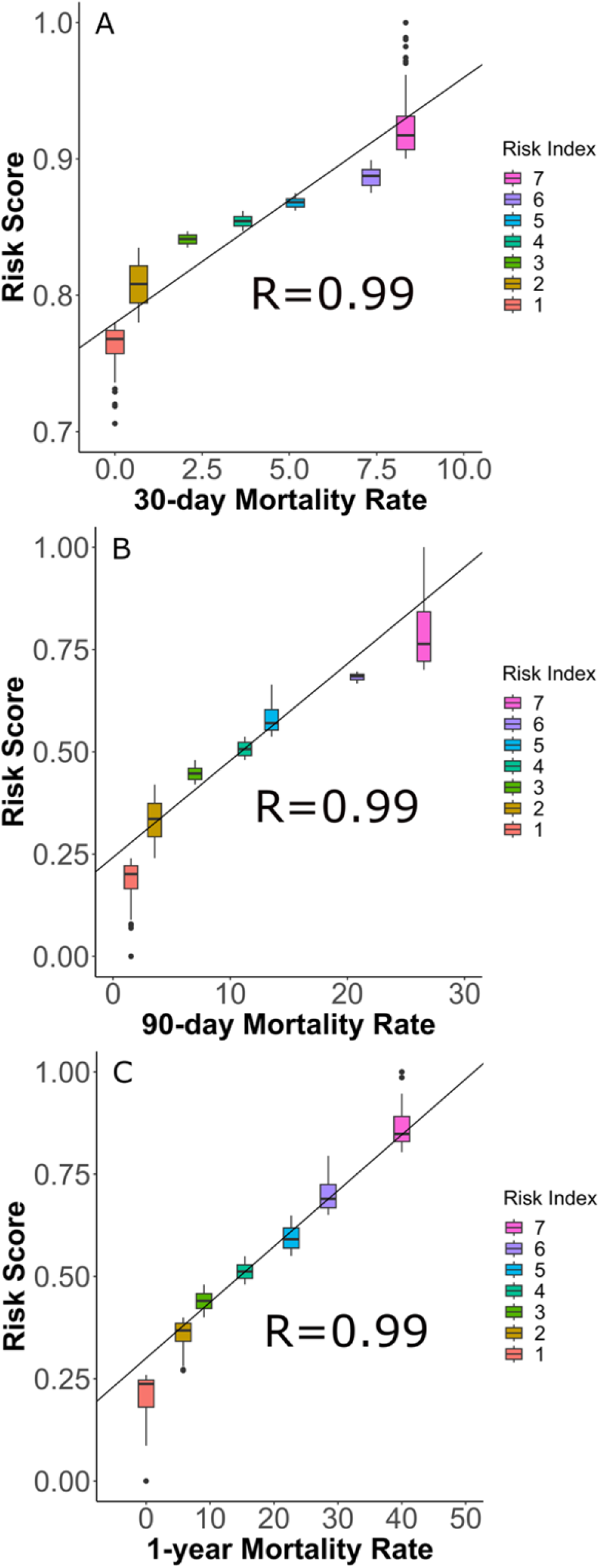
Calibration Plots for 30-day, 90-day, and 1-year Models. Patient risk scores (PRS) were calculated for all patients in the discovery cohort (N=1,179) and are provided as a function of observed patient mortality rate per tier for the 30-day, 90-day, and 1-year models. 30-day, 90-day, and 1-year models are labeled A, B, and C, respectively. The goodness-of-fit (R) was calculated using linear regression and was greater than 0.99 for all three models.

### 3.5 Logistic Regression for Predicting Short-term Mortality Outcomes

The 5-fold cross-validation procedure produced an AUC of 94.8%, 86.7.%, and 74.2% for the 30D, 90D, and 1Y additive models, respectively (**Supplemental Figures 6-8**). Using interaction terms, the AUC was 96.4%, 90.4%, and 78%, respectively (**Figure 3**). Sample hold-out validation produced an AUC of 93.8%, 92.5%, and 74.7% for the 30D, 90D, and 1Y additive models, respectively (**Supplemental Figures 9-11**). In summary, these models and the tiered risk system provide a reliable and highly accurate methodology for ranking the short-term survival of waitlisted heart transplant patients.

**Figure 3.**
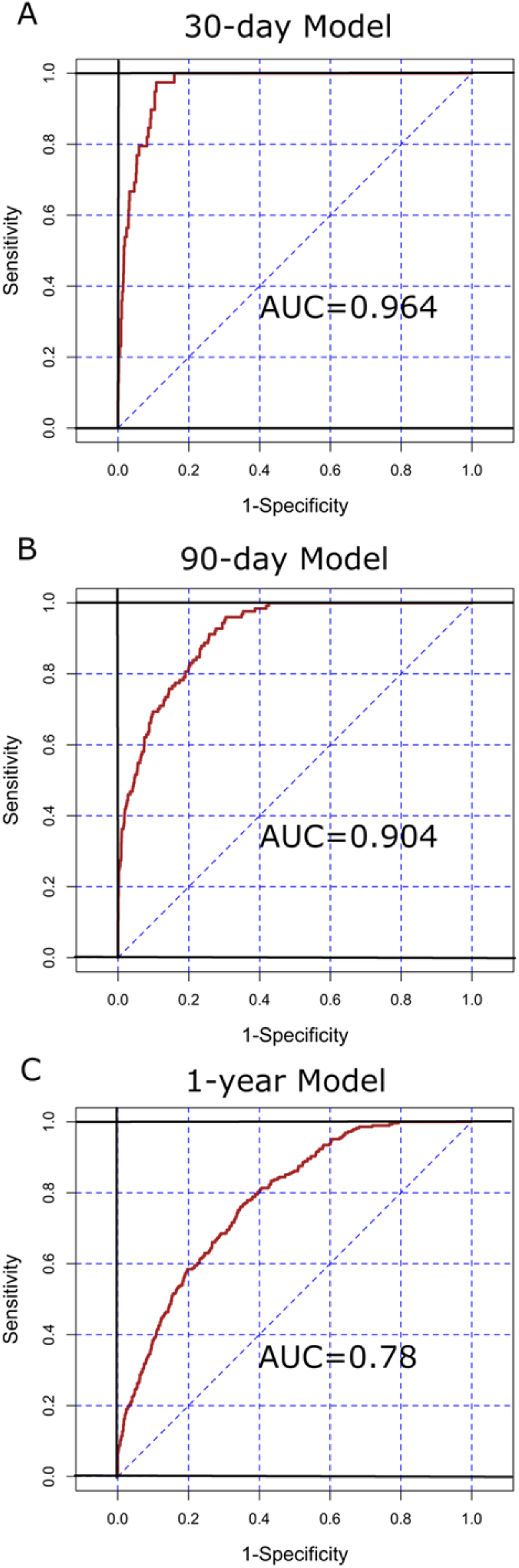
Area Under the Receiver Operating Characteristic Curve using Interaction Terms. Logistic regression was performed using the generalized linear model with interaction terms for the 30-day (A), 90-day (B), and 1-year (C) models. The area under the receiver operating characteristic curve or AUC was reported at 0.964, 0.904, and 0.78 for the 30-day, 90-day, and 1-year models, respectively.

### 3.6 Validation of the Seven-Tiered Risk Index System

Using the validation cohort, we found that the PPV of the seven-tiered risk index system ranged from 98.3% to 100% (30D), 83.7% to 100% (90D), and 0.668% to 100% (**Table 2**). The mean PPVs were 99.2% (30D), 94.1% (90D), and 88% (1Y). Using the Cox proportional-hazard regression methodology as a univariate test where the unit of measurement was the risk index, we found a significant difference in survival times by risk tier (**Supplemental Figure 12-14)**. For example, CPHR revealed that a patient with a RI of 7 had a 26% chance of death after 90 days on the waitlist, while a patient with a RI of 1 had about a 2% risk of death. The mortality rates for the risk indices provided in **Figure 2** are nearly identical to the inverse survival rates provided by the CPHR analysis.

**Table 2.**
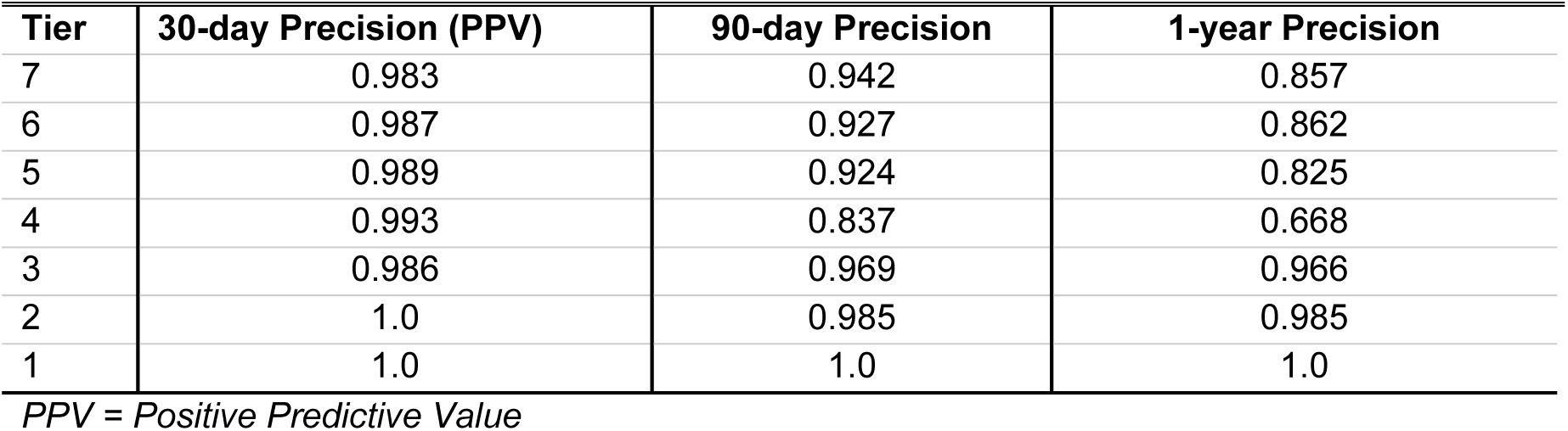
Tier Risk Index Precision for 30-day, 90-day, and 1-year Mortality Models.

| **Tier** | **30-day Precision (PPV)** | **90-day Precision** | **1-year Precision** |
| --- | --- | --- | --- |
| 7 | 0.983 | 0.942 | 0.857 |
| 6 | 0.987 | 0.927 | 0.862 |
| 5 | 0.989 | 0.924 | 0.825 |
| 4 | 0.993 | 0.837 | 0.668 |
| 3 | 0.986 | 0.969 | 0.966 |
| 2 | 1.0 | 0.985 | 0.985 |
| 1 | 1.0 | 1.0 | 1.0 |

*PPV = Positive Predictive Value*

## 4. DISCUSSION

There is currently no model that accurately stratifies waitlisted cardiac transplant patients based on medical urgency. Formal pre-transplant predictive models have been successfully developed in liver transplantation. Evans *et al*. demonstrated an overall 1-year survival rate increase of 18% in high-acuity patients in the 15 years following the national implementation of the MELD-Na score^23^. A similar metric is needed for cardiac transplant to prioritize the most critically ill waitlisted patients.

To this end, we created the Colorado Heart failure Acuity Risk Model or CHARM score. The models provided herein precisely rank patient subgroups based on waitlist mortality with mean PPVs of 99.2% (30D), 94.1% (90D), and 88% (1Y). They demonstrate excellent accuracy with AUC values of 96.4%, 90.4%, and 78%, respectively. While previous scores or models have aimed to assess heart failure illness severity, the CHARM score is the first to present this level of accuracy and predictive performance among waitlisted HF patients.

The CHARM score is intended to be used in cardiac transplant candidates at the time of listing. The 90D time frame will allow for frequent reevaluation while maintaining a high degree of prediction accuracy. The CHARM score can substantially improve heart allocation within the current “status”-based system by providing an objective prognostic measurement of medical urgency. The CHARM score will inform the continuous distribution for CD system, as it utilizes a framework that is point-based rather than “status”-based, in which candidates are prioritized for transplant through designation of a composite score from a variety of attributes. Staged implementation of the CD system is currently underway with anticipated completion of the heart allocation system within the next few years. A substantial portion (25%) of the lung transplant composite score is comprised of estimated waitlist mortality, further emphasizing the importance of pre-transplant mortality estimates for assessing patient acuity. The CHARM score provides a simple, accurate measure of pre-transplant mortality that can easily be incorporated into a heart transplant composite score.

The twelve independent patient variables incorporated into the CHARM score were chosen for their objectivity, clinical availability, and relevance to cardiac illness. Each variable was determined to significantly contribute to the predictive value of waitlist mortality. Laboratory values were selected for their evaluation of crucial organ function in the setting of severe HF. A large meta-analysis of sixty-four models that predicted death or hospitalization from HF determined renal function is one of the most significant factors in these outcomes^24^. Renal function was included in the CHARM score through serum sodium and creatinine. Multiple studies have demonstrated worsened short-term mortality for HF patients related to low serum sodium^25,26^. Creatinine is a standard measure of renal function, often used as a surrogate for eGFR. While eGFR was considered as a measure of renal function, it is not a value directly recorded in the SRTR database and can be calculated differently by institutions. Both age and BNP are also known prognostic factors in severe HF patients^17^. While age influences a variety of physiological processes, advanced age in HF (>65 years) affects vascular resistance and heart rate responsiveness, likely due to increased circulating norepinephrine levels^27^. BNP is a biomarker exclusively produced by cardiac tissue. It serves as an objective marker of cardiac stretch since it is influenced by level of end-diastolic volume, and it can also be an indicator of responsiveness to diuretic management^28^.

In addition to physiologic data, indicator variables such as previous cardiac surgery, previous cardiac transplant, ventilator support, and life support proved to be large contributors to pre-transplant mortality. These interventions often serve as a “bridge” to transplant, reserved for the most critically ill patients. Previous cardiac surgeries or transplants are also indicative of a more extensive history of cardiac illness. We specifically chose not to include variables that depend upon a physician’s practice or measurement, such as right heart catheterization data or the level of inotropic support. These variables were excluded with the intent of reducing bias based on treatment variation.

Though we present accurate predictive models, they must be viewed through the confines of the study limitations, including sample size and missing data. Sample size was dependent on the availability of data within the SRTR, reduced to 1,965 participants to account for missing laboratory variables. A thorough missingness analysis was performed to account for selection biases; however, a larger population with complete laboratory data would increase the power of these models. Additionally, variables chosen for the model were limited by the type of data recorded within SRTR. As such, certain serum markers were not readily available within the database and therefore could not be used. Similarly, the specificity of some variables was limited. For example, the term “Life Support” is broad and included anyone on inotropic support alone or varying degrees of temporary or durable mechanical support. Therefore, “Life Support” currently serves as a binary variable within the model, understanding that different modalities of mechanical support may contribute differently to patient risk. The next steps for further utilization of the CHARM score will include simulation modeling and a prospective, multi-center validation study in which these additional variables are collected and analyzed.

## Conclusion

The Colorado Heart failure Acuity Risk Model (CHARM) score provides a novel, validated model with strong positive predictive value for short-term mortality among patients waitlisted for cardiac transplantation. We anticipate the CHARM score will be useful in the era of continuous distribution to standardize organ allocation by providing an objective and intuitive system for stratifying waitlisted heart failure patients based on medical urgency.

### Author Contributions

John Stephen Malamon (JSM) had full access to all the data in the study and takes responsibility for the integrity of the data and the accuracy of the data analysis.

*Concept and design: JSM*

*Acquisition and analysis of data: JSM*

*Interpretation of data:* All authors.

*Drafting of the manuscript: RDM, SYP, JRHH, JSM*

*Critical revision of the manuscript:* All authors.

*Statistical analysis: JSM Obtained funding: BK, JRHH*

*Administrative, technical, or material support: RDM, JSM*

*Supervision: BK, JRHH, JSM*

## Data Availability

All data are obtained for publich registries.

## ACKNOWLEDGMENT

The authors have not acknowledgment to make.

